# Genetic and environmental factors underlying parallel changes in body mass index and alcohol consumption: a 36-year longitudinal study of adult twins

**DOI:** 10.1101/2022.04.08.22273585

**Authors:** Gabin Drouard, Karri Silventoinen, Antti Latvala, Jaakko Kaprio

**Affiliations:** Institute for Molecular Medicine Finland (FIMM), HiLIFE, University of Helsinki, Helsinki, Finland; Population Research Unit, Faculty of Social Sciences, University of Helsinki, Helsinki, Finland; Department of Public Health, Faculty of Medicine, University of Helsinki, Helsinki, Finland; Institute of Criminology and Legal Policy, University of Helsinki, Helsinki, Finland

**Author notes:** **Corresponding author:** Gabin Drouard, **email address:**.

**Keywords:** longitudinal twin study, genetic correlations, genetic and environmental component covariations, trajectories, heritability, baseline and changes, obesity, body mass index, alcohol consumption, behavior

## Abstract

**Background:** The genetic and environmental underpinnings of simultaneous changes in weight and alcohol consumption are poorly known.

**Objective:** We sought to quantify the environmental and genetic components underlying parallel changes in weight and alcohol consumption, and to investigate potential covariations between them.

**Methods:** The analysis comprised 4461 adult participants (58% women) from the Finnish Twin Cohort. Four measures of alcohol consumption and body mass index (BMI) were available over a 36-year follow-up. Trajectories of each trait were described by growth factors, defined as intercepts (i.e., baseline) and slopes (i.e., change over follow-up), using Latent Growth Curve Modeling. Growth values were used for male (190 MZ pairs, 293 DZ pairs) and female (316 MZ pairs, 487 DZ pairs) same-sex complete twin pairs in multivariate twin modeling. The variance and covariance of growth factors were then decomposed into genetic and environmental components.

**Results:** The baseline heritabilities were similar in men (BMI: h^2^=79%; alcohol consumption: h^2^=49%) and women (h^2^=77%; h^2^=45%). Heritabilities of BMI change were similar in men (h^2^=52%) and women (h^2^=57%), but higher in men for change in alcohol consumption (h^2^=45%) than in women (h^2^=31%). Significant genetic correlations between BMI at baseline and change in alcohol consumption were observed in both men (r =-0.17(95% Confidence Interval: -0.29,-0.04)) and women (r=-0.18(−0.31,-0.06)). The genetic components of baseline and longitudinal change were correlated for both BMI and alcohol consumption with sex differences. Non-shared environmental factors affecting changes in alcohol consumption and BMI were correlated in men (r=0.18(0.06,0.30)). Among women, non-shared environmental factors affecting baseline alcohol consumption and the change in BMI were correlated (r=-0.11(−0.20,-0.01)).

**Conclusions:** We provide evidence of genetic correlations between BMI and change in alcohol consumption. Independent of genetic effects, change in BMI and change in alcohol consumption covary.

## Introduction

Obesity and alcohol use are major public health issues worldwide due to their high and increasing prevalence in recent decades (1-2). Body mass index (BMI) is a widely used indicator for measuring obesity and is associated with a large number of diseases such as cardiovascular diseases, several cancers and mental health (3), reinforcing the value of studying the genetics of both obesity and weight change for preventive purposes. The genetics of obesity is well-established through twin and family studies (4) and molecular genetic studies (5-6), paving the way for both precision nutrition (7) and nutrigenomics (8).

Alcohol consumption, despite its high caloric content (9), influences weight gain in a way that remains unclear (10) due notably to nutritional disorders induced by alcohol use (11). The relationship between obesity and alcohol-related behaviors hinges on plural and complex patterns incorporating dimensions of mental health (3,12), behavioral sciences (13), and molecular biology (14). A weak genetic correlation between BMI and drinks per day has also been recently reported and was estimated to be -0.08 in a very large multicohort genome-wide association study (GWAS) (15).

Alcohol use is widely known to have a genetic component (16-17) and substantial interindividual differences, such as sex differences (18-20). Large epidemiological studies have also found differences and inconsistencies between men and women in the association between drinking at baseline and weight gain (21-24). These findings encourage a sex-specific approach to the study of the relationship between BMI and alcohol consumption. Large-scale studies of how BMI predicts changes in alcohol use appear to be rare.

The aims of the current study are 1) to study the longitudinal relationship between BMI and alcohol consumption trajectories, 2) to highlight the genetic and environmental components underlying changes in BMI and alcohol consumption, and 3) to investigate possible environmental or genetic structures underlying BMI and alcohol consumption trajectories. To this end, we derived indicators of BMI and alcohol consumption trajectories using Latent Growth Curve Modeling (LGCM) from a cohort of twins followed over 36 years. Genetic and environmental influences associated with changes in BMI and alcohol consumption and their mutual associations were then estimated using multivariate twin modeling. Further, we investigated sex differences in these associations.

## Materials and Methods

### Participants

Data from the older Finnish Twin Cohort (FTC) were used in the current study. The FTC is a large-scale, population-based twin cohort established in 1974 (25), which provides valuable resources for the study of complex phenotypes, such as obesity and alcohol use (26-27). The study participants were same-sex monozygotic (MZ) and dizygotic (DZ) twins born before 1958 (28), for which questionnaire data were first collected in 1975. FTC participants responded to up to four waves of questionnaires from surveys conducted in 1975, 1981, 1990, and 2011. The data included measures of weight, height and alcohol consumption. Self-reported height and weight measures were found to be reliable when they were examined from a subsample of participants who completed the 2011 questionnaire: correlations between measured and self-reported BMI were 0.95 for both men and women (29).

A total of 24,384 adult same-sex twin participants had data from at least one survey, and they entered the data preprocessing phase (Figure 1). After applying exclusion criteria, 4461 participants constituted the final sample (58% females). The male subsample included 483 complete same-sex twin pairs (34% MZ pairs), and the female subsample included 803 complete same-sex twin pairs (40% MZ pairs). The pre-processing steps required to obtain the final sample are contextualized in Figure 1 and detailed as follows:

**Figure 1:**
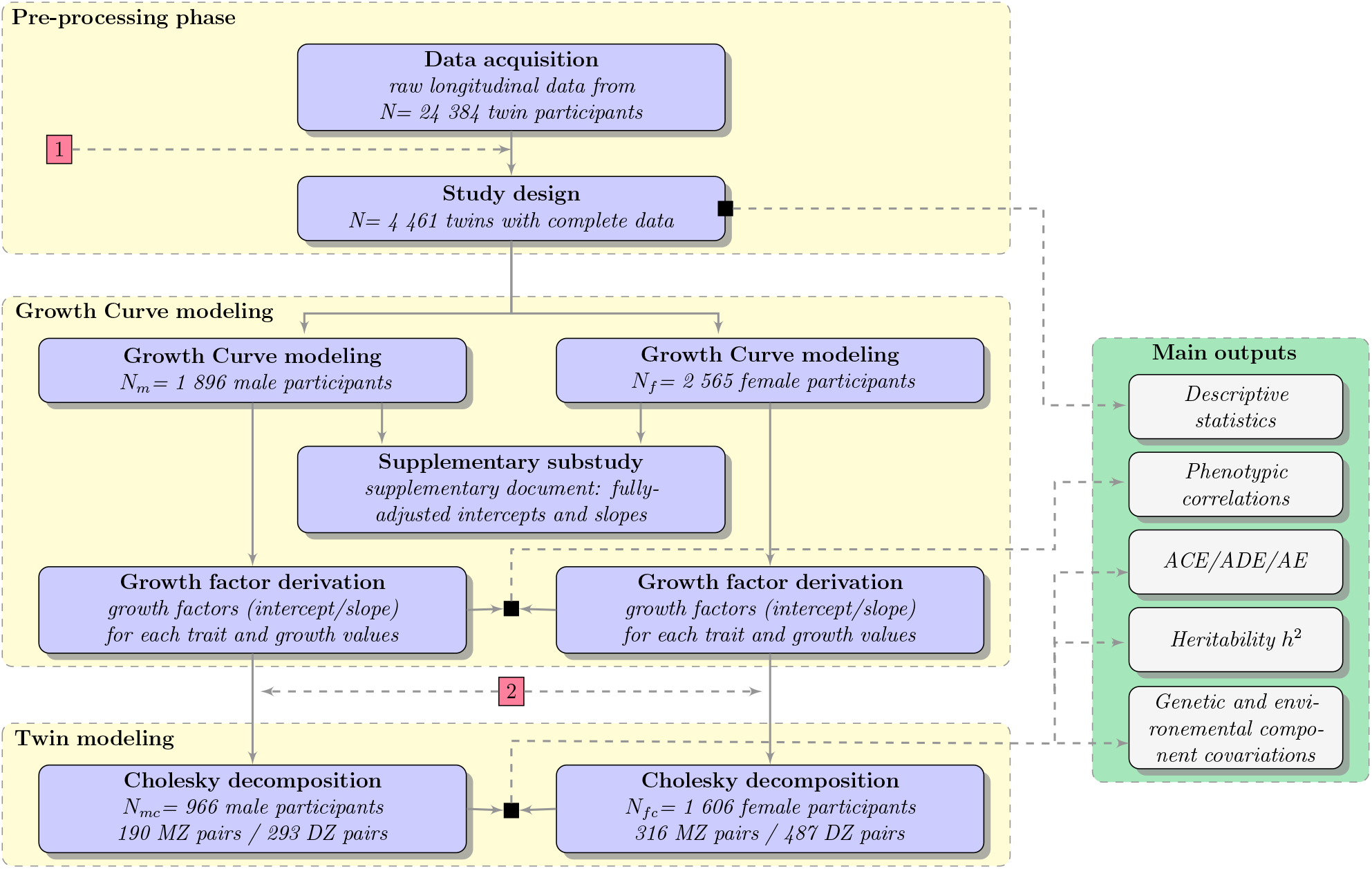
Title: General study plan diagram legend: *The study was structured in two main models based on latent representations, following a data preprocessing phase*. Sex-specific growth factors were derived to study trajectories and phenotypic correlations at the population level. The Twin modeling was based on growth values derived from the LGCM. Two exclusion criteria were performed at different times and are represented by pink squares: they correspond to (1) data preprocessing and (2) selection of complete same-sex twin pairs. MZ: monozygotic twins. DZ: dizygotic twins. N: number of participants. LGCM: Latent Growth Curve Modeling.

1. BMI measures for each wave were calculated using the associated participant height and weight measures. Height measurements identified as missing were supplemented with height measurements from other questionnaires when the information was available. Three participants with inconsistent BMI values were excluded from the study. Alcohol measures, expressed in grams per month (17), were log-transformed as well as BMI measures to remove the skewness of the original variable (BMI skewness ranged 0.62–1.07 in men and 1.07–2.47 in women before log-transformation; 0.25–0.48 and 0.48–1.11, respectively, after) (16,30). To correct for any zeros that may be present in the alcohol consumption variable, *f*: *t* ↦ *log*(1 + *t*) was used as log-transformation for the alcohol consumption measures. Of the initial participants, only those whose BMI and alcohol measures were complete on all four questionnaires were retained. This pre-selection led to a first sample of 4663 participants (Figure 1).
2. The use of covariates for additional exploratory purposes was considered (31-33) (Supplementary Document, Table S1, Table S2), and led to a reduction of the sample. Among these covariates, a variable indicating smoking status was not recorded for 202 participants. Despite the absence of these covariates in our study, participants with missing smoking data were excluded to compare the phenotypic correlations obtained in the main study and those produced in the supplementary sensitivity analysis (Supplementary Document, Table S3). This exclusion criterion led to a final sample of 4461 participants.

Overall, the average weight and alcohol consumption among male and female participants tended to increase over time. Alcohol consumption in men ranged from 2.29-fold to 2.56-fold more than women across the 4 questionnaires. The average BMI increased over the period 1975–2011 from 22.5 (standard deviation: 2.5) kg/m^2^ to 26.6 (3.9) in men, and from 20.8 (2.5) to 25.7 (4.4) in women. The distributions of logarithmic BMI, logarithmic alcohol consumption, age, and untransformed raw values across questionnaires are described in Table 1. Multivariate twin modeling was performed only on the subset of complete same-sex pairs.

**Table 1:** Title: Descriptive statistics of age, BMI, alcohol consumption, and growth factor estimates in men and women across the four questionnaires. legend: Indicators of age, log-BMI, BMI, log-alcohol consumption and alcohol consumption distributions are summarized by the mean and standard deviation. Growth factors are summarized as population-wide means, provided with 95% confidence intervals. More information on growth factor distributions can be found in the Supplementary Document. ALC: logarithmic alcohol consumption. CI: confidence interval; sd: standard deviation.

|  |  | BMI (kg/m<sup>2</sup>) |  |  |  | ALC (g/month) |  |  |  | Age (years) |  |  |  |
| --- | --- | --- | --- | --- | --- | --- | --- | --- | --- | --- | --- | --- | --- |
|  |  | Men |  | Women |  | Men |  | Women |  | Men |  | Women |  |
| raw values | questionnaire | mean | sd | mean | sd | mean | sd | mean | sd | mean | sd | mean | sd |
|  | 1975 | 22.49 | 2.5 | 20.83 | 2.5 | 339.38 | 397.0 | 143.23 | 187.2 | 24.04 | 3.7 | 23.57 | 3.7 |
|  | 1981 | 23.49 | 2.7 | 21.62 | 2.8 | 341.83 | 393.3 | 136.6 | 188.9 | 30.28 | 3.7 | 29.81 | 3.7 |
|  | 1990 | 24.73 | 3.1 | 23.09 | 3.7 | 419.90 | 477.4 | 182.89 | 255.7 | 39.35 | 3.7 | 38.86 | 3.7 |
|  | 2011 | 26.62 | 3.9 | 25.72 | 4.4 | 508.48 | 678.7 | 198.12 | 307.3 | 60.45 | 3.7 | 59.94 | 3.7 |
| log values | questionnaire | mean | sd | mean | sd | mean | sd | mean | sd |  |  |  |  |
|  | 1975 | 3.11 | 0.1 | 03.03 | 0.1 | 05.03 | 1.7 | 4.19 | 1.7 |  |  |  |  |
|  | 1981 | 3.15 | 0.1 | 03.07 | 0.1 | 5.14 | 1.6 | 4.10 | 1.7 |  |  |  |  |
|  | 1990 | 3.20 | 0.1 | 3.13 | 0.1 | 5.36 | 1.5 | 4.38 | 1.7 |  |  |  |  |
|  | 2011 | 3.27 | 0.1 | 3.23 | 0.2 | 5.46 | 1.6 | 4.39 | 1.7 |  |  |  |  |
|  |  | mean | CI | mean | CI | mean | CI | mean | CI |  |  |  |  |
| growth values | Intercept | 3.12 | (3.12,3.13) | 03.03 | (3.03,3.04) | 05.09 | (5.02,5.16) | 4.16 | (4.10,4.22) |  |  |  |  |
|  | Slope | 0.004 | (0.004,0.005) | 0.006 | (0.006,0.006) | 0.011 | (0.009,0.014) | 0.007 | (0.005,0.009) |  |  |  |  |

### Latent Growth Curve Modeling

Structural Equation Modeling (SEM) has become a common set of methods used in the analysis of behavioral phenotypes (34-35). We used LGCM, an SEM-based method, to represent repeated measures data as a function of time. The linear LGCM derives two latent variables, hereafter called growth factors, to characterize within-person change over time. The growth factors consist of an intercept and a slope that can be interpreted as a rate of change. The means of the growth factors therefore describe the average trajectory at the population level, while their (co-)variances provide key information about the magnitude of inter-individual differences.

The LGCM, in the study of parallel processes, allows 1) intercepts and slopes of traits to freely covary or 2) estimation of growth factors to be constrained from other growth factors. In our multivariate study, the slopes and intercepts derived from the four measures of BMI and alcohol consumption were freely derived. That is, they were free to correlate without influencing each other during the model-fitting phase (Figure 2). This modeling was sex-specific to correct for structural differences in alcohol consumption between the sexes that are well-established in most populations (16,18-20). Nullity tests of the means and variances of the growth factors were performed to describe the trajectory of each trait, for each sex. Phenotypic correlations between growth factors were also calculated and growth values were used in twin modeling (Figure 1). A p-value lower than 0.05 was considered sufficient to reject the null hypothesis of nullity of the mean, variance and correlation. The modeling was conducted using the R package lavaan (36), version 0.6-10.

**Figure 2:**
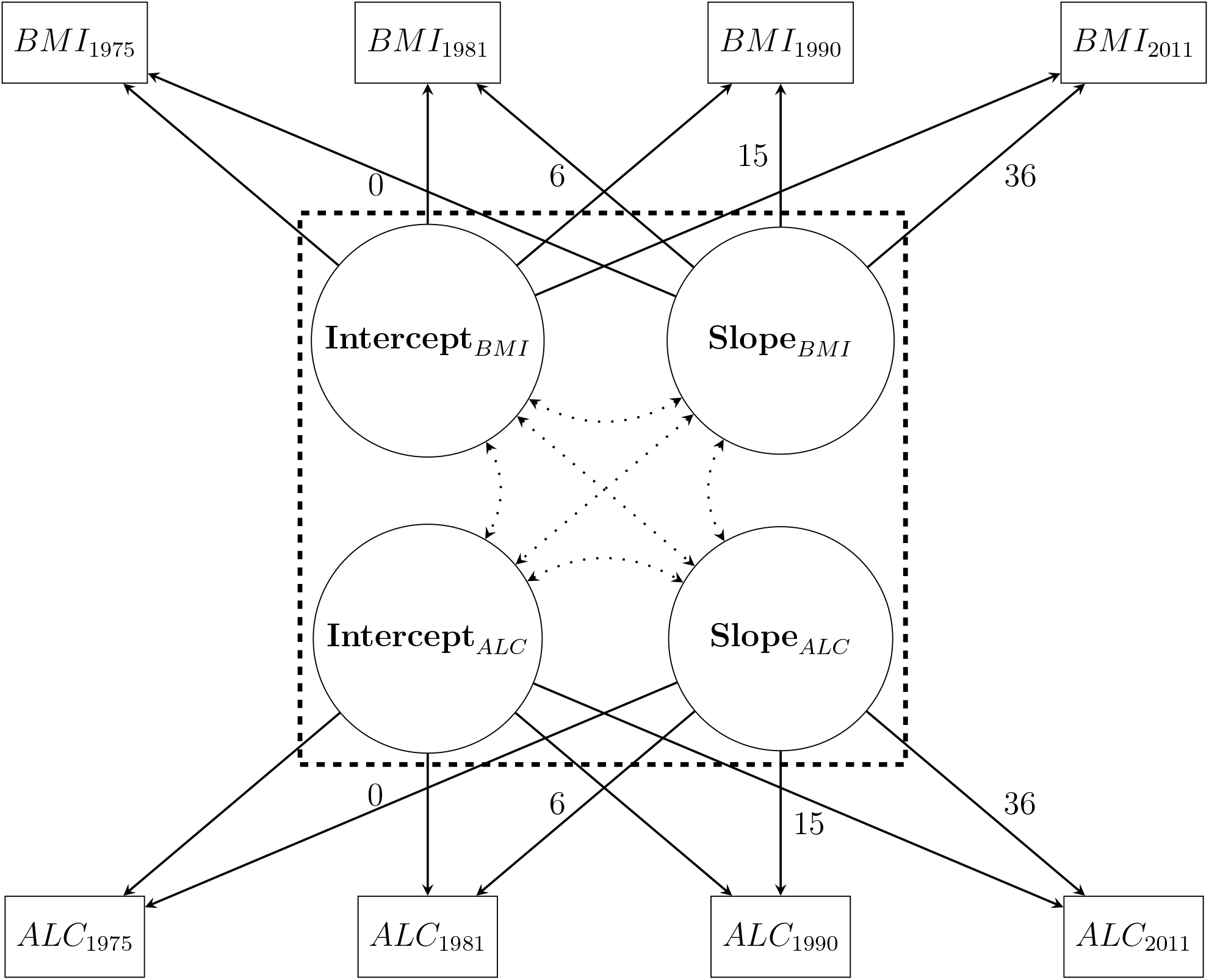
Title: Diagram of Latent Growth Curve Modeling used to study participants’ BMI and alcohol consumption sex-specific trajectories. legend: *Growth factors characterizing individual trajectories were derived for each trait, for males and females separately*. Logarithmic measures of BMI (*BMI* _1975_, *BMI* _1981_, *BMI* _1990_, *BMI* _2011_) and alcohol consumption (*ALC*^*^_1975_, *ALC*^*^_1981_, *ALC*^*^_1990_, *ALC*^*^_2011_) were used to derive growth factors (intercept and slope). The dashed box is the targeted latent representation underlying the parallel trajectories of log-BMI and log-alcohol consumption. The growth factors were freely covaried in the model fitting phase (dotted two-way arrows).

### Multivariate twin modeling

The epidemiological value of using twin cohorts to understand complex phenotypes is well established (26). One of the applications of twin study designs is quantitative genetic modeling, based on the comparison of MZ and DZ twins. MZ twins share 100% of their DNA, while DZ twins share on average 50% of their segregating genes. This model sources on the differences in phenotypic intra-class correlations between MZ and DZ twins: these are assumed to reflect genetic and environmental components.

Classical twin modeling decomposes the total trait variance into genetic and environmental components. If the DZ twin intra-class correlation (rDZ) is less than half of the MZ twin intra-class correlation (rMZ), the genetic component can be subdivided into an additive (A) and non-additive (D) component, reflecting the summed allelic effects across genes and the genetic dominance, respectively. If rDZ is greater than half of rMZ, a shared (C) environmental component reflecting shared environmental effects (e.g., such as family common environment and other effects that make siblings similar) can be estimated. Further, in all models, a non-shared (E) environmental component (i.e., unique to each sibling) will be estimated reflecting individual-level environmental factors as well as measurement error. Since the D and C components cannot be estimated simultaneously, the combinations of classical twin modeling in a univariate context where a genetic component is considered certain are mainly summarized in the ACE and ADE models, which can be compared downstream to a simplified AE model.

We calculated the intraclass correlations of growth factors between DZ and MZ twins to determine the most appropriate C or D component for model fitting (Table 2). The pattern of correlation for the alcohol consumption intercept suggested the estimation of the C component, rather than the D component: the ratio of DZ to MZ intraclass correlations was substantially greater than 50% in women and men. The set of intra-class correlation estimates with their associated 95% confidence intervals is provided in Table 2.

**Table 2:** Title: Phenotypic correlations between growth factors and intra-class correlations in male and female participants. legend: Phenotypic correlation coefficients are associated with 95% confidence intervals. Intra-class correlations for MZ and DZ twins are associated with 95% confidence intervals. The phenotypic correlation coefficients for which the null hypothesis of nullity cannot be rejected at the 5% level are in italics. Phenotypic correlation coefficients for which the confidence intervals do not cross those of the opposite sex are highlighted in bold. ALC: alcohol log-consumption. MZ: monozygotic. DZ: dizygotic. M: Men. W: Women.

|  |  | Phenotypic correlations |  |  |  | Intra-class correlations |  |
| --- | --- | --- | --- | --- | --- | --- | --- |
|  |  | Intercept BMI | Slope BMI | Intercept ALC | Slope ALC | MZ | DZ |
| M | Intercept BMI | 1 |  |  |  | 0.79<br>(0.73, 0.84) | 0.45<br>(0.36, 0.54) |
|  | Slope BMI | **-0.10**<br>**(-0.14, -0.05)** | 1 |  |  | 0.51<br>(0.40, 0.61) | 0.24<br>(0.13, 0.34) |
|  | Intercept ALC | **0.15**<br>**(0.11, 0.20)** | -0.05<br>(-0.09, -0.00) | 1 |  | 0.71<br>(0.63, 0.77) | 0.47<br>(0.38, 0.56) |
|  | Slope ALC | **-0.19**<br>**(-0.23, -0.15)** | **0.16**<br>**(0.11, 0.20)** | **-0.55**<br>**(-0.58, -0.51)** | 1 | 0.47<br>(0.35, 0.57) | 0.25<br>(0.14, 0.35) |
| W | Intercept BMI | 1 |  |  |  | 0.79<br>(0.74, 0.83) | 0.34<br>(0.26, 0.42) |
|  | Slope BMI | **0.08**<br>**(0.04, 0.12)** | 1 |  |  | 0.57<br>(0.49, 0.64) | 0.31<br>(0.22, 0.38) |
|  | Intercept ALC | **-0.02**<br>**(-0.06, 0.02)** | -0.06<br>(-0.10, -0.02) | 1 |  | 0.64<br>(0.57, 0.70) | 0.46<br>(0.38, 0.52) |
|  | Slope ALC | **-0.09**<br>**(-0.13, -0.05)** | **0.02**<br>**(-0.01, 0.06)** | **-0.33**<br>**(-0.37, -0.30)** | 1 | 0.36<br>(0.26, 0.45) | 0.11<br>(0.02, 0.20) |

The heritability h^2^ is defined, in this classical configuration, as the share of the phenotypic variance attributed to additive genetic variance. The coefficients e^2^ and c^2^ (or d^2^) are defined as above and provide information on the part of the variance explained by the individual’s unique environment, and the environment shared by the two co-twins (or genetic dominance, respectively). In a multivariate context, the covariances between traits can also be decomposed using the Cholesky decomposition. The genetic and environmental components of each trait can then be correlated. The path coefficients associated with the respective components A, E, and C or D thus provide information on the cross influence of genetic and environmental components on the set of growth factors.

We investigated the genetic and environmental influence of the growth factors of BMI and alcohol consumption in a multivariate setting. Twin modeling was performed using the complete same-sex twin pairs derived from the LGCM with their growth values (Figure 1). Because of heterogeneous intra-class correlation between MZ and DZ twins within the four latent variables (Table 2), several models were tested. An unconstrained saturated model estimating all parameters was first performed to build models based on A, E and C or D components to ensure that the twin modeling assumptions were met. These assumptions include the equality of means and variances between co-twins and between zygosity types (37): we therefore tested these assumptions, for each sex. The equality of environments between MZ and DZ twins was also assumed. Evaluation of the ACE and ADE models was investigated by comparing them to the AE model through two performance measures: the likelihood with the -2 log-likelihood measure (−2LL) and the Akaike information criterion (AIC), which must be maximized and minimized, respectively (38). The AIC criterion was preferred to the likelihood criterion when neither showed the superiority of one model over another. Confidence intervals for the parameters (h^2^, e^2^, c^2^ and d^2^) and for correlations between genetic components and between environmental components were calculated with a 95% confidence margin. The genetic twin modeling was conducted using the R package OpenMx (39), version 2.20.3.

An ACE and ADE model were fitted for men and women by decomposing the variation of each growth factor into A, C or D, and E variance components. These two models, hereafter referred to as full ACE and full ADE, showed significant improvement in model fitting compared with the saturated model in both men and women (p-value < 0.05, likelihood ratio test). These two models were then compared with the AE model, deriving only an additive genetic component and a non-shared environmental component. The AE model outperformed the full ADE and full ACE models, both in terms of AIC and log-likelihood, for modeling of men (Table 3). The full ACE and full ADE models showed better AIC performance than the AE model (full ACE: -20161.51 vs. AE: -20160.96; full ADE: -20159.51 vs AE: -20158.96) for women, but not for men. The log-likelihood of the AE model was better than that of the full ACE and full ADE models in both sexes. To remove the inflection effect of the shared environmental component estimate for all growth factors on the performance measures, a partial ACE model deriving only a shared environmental component at the alcohol intercept was constructed because of 1) the intraclass correlation ratio of this growth factor for both sexes and 2) the significant c^2^ component estimate in the full ACE model (Table 3). This partial model increased the difference in AIC with the AE model compared to the differences observed with the full ACE model. The partial ACE model also outperformed the ADE model when compared to the saturated model, both in terms of likelihood (partial ACE: -2LL = -12498.52; ADE: -2LL = -12508.55) and AIC (partial ACE: -12448.52; ADE: -12438.55). This model being the most parsimonious, it was kept as the final model.

**Table 3:** Title: Estimation of heritability and environmental parameters of multivariate AE, ACE, and ADE models and their fit indices. legend: Models that fit significantly better than the AE model have their performance measures highlighted in bold. The parameter estimates are provided with a 95% confidence interval in the form *estimate (lower bound, higher bound)*. **\*/** refers to unreliable measures, i.e., growth factors with an estimated C or D component while the ratio of intra-class correlations (Table 2) prohibits their estimation. Non-significant estimates are in italics. -2LL: -2 log-likelihood. AIC: Akaike information criterion. ALC: alcohol log-consumption.

| Twin Model | Growth factor | Men |  |  |  | Women |  |  |  |
| --- | --- | --- | --- | --- | --- | --- | --- | --- | --- |
|  |  | AIC & -2LL<br>(vs AE) | A | C or D | E | AIC & -2LL<br>(vs AE) | A | C or D | E |
| AE | Intercept BMI | NA | 79% (74,83) | *set to 0%* | 21% (17,26) | NA | 77% (73,81) | *set to 0%* | 23% (19,27) |
|  | Intercept ALC |  | 70% (64,75) | *set to 0%* | 30% (25,36) |  | 65% (60,70) | *set to 0%* | 35% (30,40) |
|  | Slope BMI |  | 52% (42,61) | *set to 0%* | 48% (39,58) |  | 57% (50,63) | *set to 0%* | 37% (37,50) |
|  | Slope ALC |  | 45% (35,54) | *set to 0%* | 55% (46,65) |  | 31% (22,38) | *set to 0%* | 69% (62,78) |
| full ACE | Intercept BMI | AIC:<br>-12440.55<br>(vs -12444.25) | 63% (46,78) | 16% (2,31) | 21% (17,27) | AIC:<br>**-20161.51**<br>(vs **-20160.96**) | 77% (67,81) | *0% (0,8)* | 23% (19,27) |
|  | Intercept ALC |  | 39% (19,60) | 29% (11,46) | 32% (26,39) |  | 30% (13,46) | 32% (18,46) | 38% (32,44) |
|  | Slope BMI | -2LL:<br>-12508.55<br>(vs -12492.25) | 40% (19,55) | *10% (0,25)* | 50% (41,60) | -2LL:<br>-20229.51<br>(vs -20208.96) | 50% (30,61) | *6% (0,22)* | 44% (38, 51) |
|  | Slope ALC |  | 33% (8,50) | *11% (0,31)* | 56% (47,67) |  | *19% (4,32) | *10% (2,22) | *71% (63,79) |
| partial ACE | Intercept BMI | AIC:<br>**-12446.78**<br>(vs **-12444.25**) | 79% (74,83) | *set to 0%* | 21% (17,26) | AIC:<br>**-20163.54**<br>(vs **-20160.96**) | 77% (73,81) | *set to 0%* | 23% (19,27) |
|  | Intercept ALC |  | 49% (32,67) | 19% (5,33) | 31% (25,38) |  | 45% (29,62) | 18% (4,32) | 37% (31,43) |
|  | Slope BMI | -2LL:<br>-12500.78<br>(vs -12492.25) | 52% (42,61) | *set to 0%* | 48% (39,58) | -2LL:<br>-20217.54<br>(vs -20208.96) | 57% (50,63) | *set to 0%* | 37% (37,50) |
|  | Slope ALC |  | 45% (34,54) | *set to 0%* | 55% (46,66) |  | 31% (22,38) | *set to 0%* | 69% (62,78) |
| full ADE | Intercept BMI | AIC:<br>-12438.55<br>(vs -12442.25) | *59% (47,76) | *24% (4,38) | *17% (12,22) | AIC:<br>**-20159.51**<br>(vs **-20158.96**) | 77% (65,81) | *0% (0,15)* | 23% (18,27) |
|  | Intercept ALC |  | *43% (34,58) | *37% (18,48) | *20% (15,27) |  | *38% (30,47) | *39% (27,48) | *23% (19,28) |
|  | Slope BMI | -2LL:<br>-12508.55<br>(vs -12492.25) | 42% (28,55) | 17% (1,34) | 41% (32,52) | -2LL:<br>-20229.51<br>(vs -20208.96) | 50% (36,61) | *11% (0,31)* | 39% (30,48) |
|  | Slope ALC |  | 36% (22,50) | *18% (0,38)* | 46% (35,60) |  | 24% (16,33) | 17% (4,31) | 59% (48,70) |

Because of differences in samples induced by the stratification of the study by sex, statistical power to detect genetic and environmental components was assessed in men. Since the average heritability for behavioral traits is generally estimated to be around 50% (40-41), we calculated the power associated with the detection of component A in an AE (or C in an ACE) model within the male twin pair subgroup if the additive component was A=50% (or A=50% and C=20%, respectively) (42-43). The power associated with the detection of A was estimated to be 95%. However, the power associated with the detection of C, when the shared environmental component is 20%, was estimated at 65.6% while it was 84.0% in women. Our design was therefore well powered in women, but some weaknesses in statistical power were observed in the male twin subgroup to detect modest shared environmental components.

## Results

### Trajectories and phenotypic correlations

Phenotypic correlation analysis was performed using growth values characterizing trait trajectories derived from the LGCM (Figure 1). The trajectory associated with log alcohol consumption showed a general trend, among both men and women, toward a generalized increase over the period 1975–2011. This rate was 0.007 log-units per year from baseline for women and 0.011 log-units per year for men from baseline. The population-wide trend in BMI increase was 0.004 log BMI units per year in men and 0.006 log BMI units per year in women (Table 1). Additional information on log BMI trajectories, log alcohol consumption trajectories and growth factor distributions can be found in the Supplementary Document.

The correlation between intercept and slope of alcohol consumption was moderate in men (r = -0.55, 95% CI: -0.58, -0.51) and somewhat weaker in women (r = -0.33, 95% CI: -0.37, -0.30). The relationship between BMI intercept and BMI slope was positive in women (r = 0.08, 95% CI: 0.04, 0.12) but negative in men (r = -0.10, CI: -0.14, -0.05). Alcohol and BMI slopes were significantly correlated and estimated at 0.16 (95% CI: 0.11, 0.20) in men, but were not significant in women (r = 0.02, 95% CI: -0.01, 0.06). Phenotypic correlations between growth factors were therefore found to be of different magnitude, sign or significance depending on sex. The set of correlations is shown in Table 2.

A sensitivity analysis was performed to compare the phenotypic correlations of the study with those obtained by adjusting log BMI and log alcohol consumption variables for covariates such as smoking status, marital status or education level in addition to the age. This comparative analysis is shown in the Supplementary Document (Supplementary Document, Figure S1).

### Multivariate twin modeling

A total of 483 same-sex pairs of male twins and 803 same-sex pairs of female twins were used to decompose the variance and covariance of the LGCM-derived growth factors (Figure 1).

The heritability of the BMI intercept was estimated to be h^2^ = 79% (95% CI: 74, 83) in men and h^2^ = 77% (95% CI: 73, 81) in women, suggesting no sex-related difference in the heritability estimate of the BMI intercept. The share of heritability within alcohol intercepts was estimated at h^2^ = 49% (95% CI: 32, 67) in men and h^2^ = 45% (95% CI: 29, 62) in women. The common environmental variance component for the alcohol consumption intercept was estimated at c^2^ = 19% (95% CI: 5, 33) in men and c^2^ = 18% (95% CI: 4, 32) in women. All the estimates of the full ACE, full ADE, partial ACE and AE models can be found in Table 3.

The genetic components of alcohol consumption and BMI slopes, however, showed notable differences by sex. Men, for whom the relationship between alcohol consumption intercept and slope was significantly stronger than in women (Table 3), were also conditioned by a stronger hereditary component for alcohol slope that was estimated to be h^2^ = 45% (95% CI: 34, 54), whereas it was h^2^ = 31% (95% CI: 22, 38) in women. On the other hand, the non-shared environment played a more important role in BMI change for men at e^2^ = 48% (95% CI: 39, 58) than for women at e^2^ = 37% (95% CI: 37, 50). The heritability of BMI change was therefore estimated to be h^2^ = 52% (95% CI: 42, 61) in men, and h^2^ = 57% (95% CI: 50, 63) in women.

A positive genetic correlation was also observed between the BMI intercept and the BMI slope in women (r = 0.24, 95% CI: 0.14, 0.33), but not in men (r = -0.08, 95% CI: -0.19, 0.05). The additive genetic components of the intercept and slope of alcohol consumption were found to be significantly correlated for women (r = -0.37, 95% CI: -0.49, -0.24) and men (r = -0.82, 95% CI: -1.00, -0.65), but with different magnitudes (Figure 3). A similar genetic correlation between BMI intercept and alcohol consumption slope was observed in men (r = -0.17, 95% CI: -0.29, -0.04) and women (r = -0.18, 95% CI: -0.31, -0.06). These results support the existence of complex genetic architectures influencing 1) alcohol consumption and changes in alcohol consumption, 2) BMI with changes in BMI, and 3) BMI intercept with alcohol-related behavior over time. No other genetic correlations between BMI and alcohol consumption growth factors reached significance.

**Figure 3:**
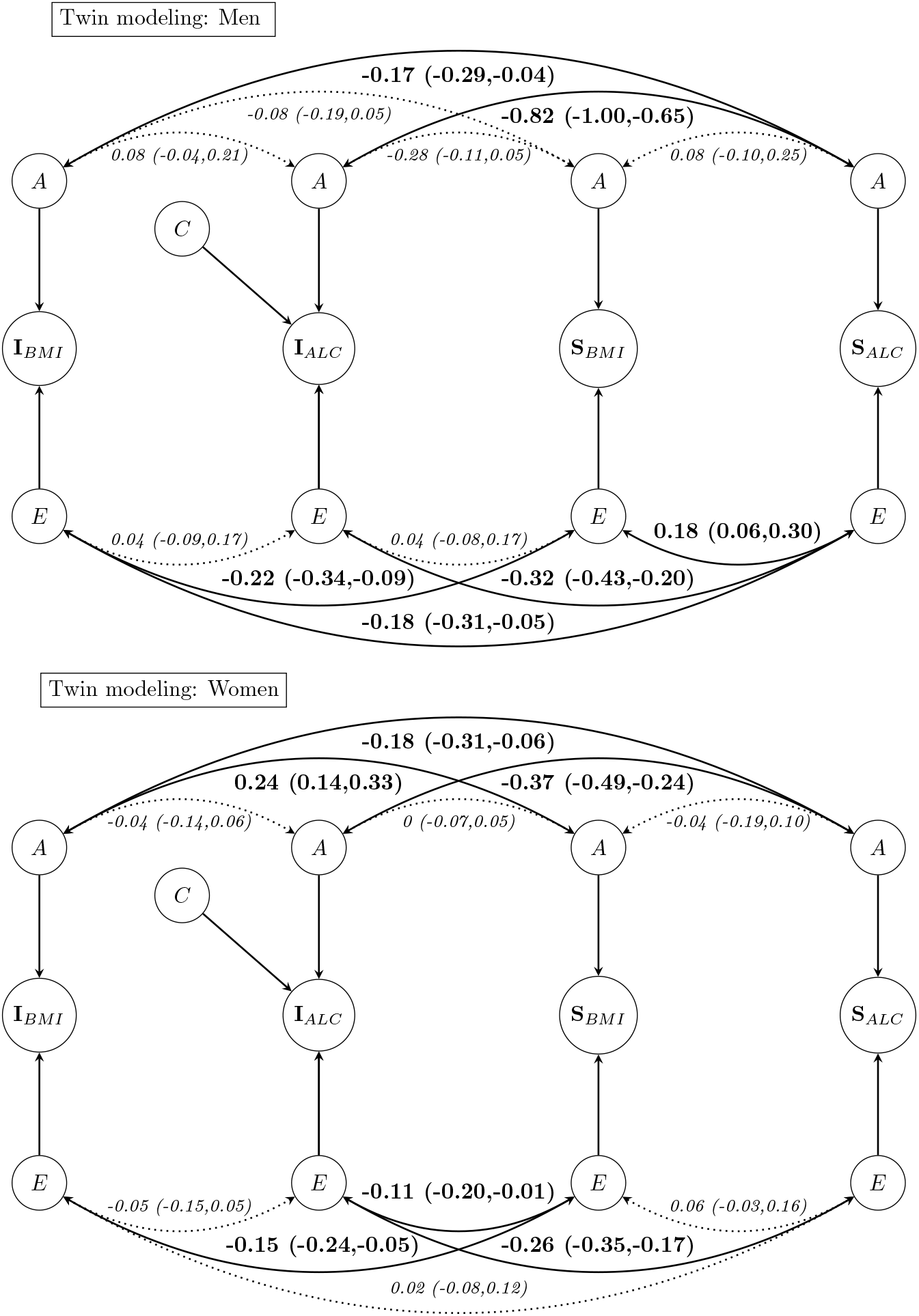
Title: Path diagrams illustrating covariations between genetic and environmental components within the best performing multivariate Twin modeling (partial ACE) for male and female participants. legend: *Within-trait covariations between genetic components highlighted a key hereditary structure in weight gain and change in alcohol consumption. Sex-specific covariations between non-shared environmental components showed a common environmental influence in BMI and alcohol consumption trajectories*. Estimates are provided with a 95% confidence interval (95% CI lower bound, 95% CI higher bound), bolded when the estimate was significant; italicized otherwise. The components are linked by solid lines when correlations were significant; dotted lines otherwise. *I* _*BMI*_ and *I* _*ALC*_ correspond to the BMI and alcohol log-consumption intercepts, respectively. *S*_*BMI*_ and *S*_*ALC*_ correspond to BMI and alcohol log-consumption slopes, respectively. A: additive genetic component. C: shared environmental component. E: non-shared environmental component. CI: confidence interval.

Correlations between non-shared environmental components were also widely observed (Figure 3). The non-shared environmental components of BMI intercepts and slopes were negatively correlated in men (r = -0.22, 95% CI: -0.34, -0.09) and women (r = -0.15, 95% CI: -0.24, -0.05). Growth factors for alcohol consumption also covaried via their environmental components in both men (r = -0.32, 95% CI: -0.43, -0.20) and women (r = -0.26, 95% CI: -0.35, -0.17). In addition to environmental correlations between growth factors of the same trait, correlations between environmental components of BMI and alcohol consumption were observed. The non-shared environmental components of alcohol consumption intercept and BMI slope were negatively correlated in women (r = -0.11, 95% CI: -0.20, -0.01). BMI change was also positively associated with alcohol consumption change in men via their non-shared environmental components (r= 0.18, 95% CI: 0.06, 0.30). All correlation estimates between environmental components are presented in Figure 3.

## Discussion

The objectives of our study were 1) to study the longitudinal relationship between BMI and alcohol consumption trajectories, 2) to highlight the genetic and environmental components underlying changes in weight and alcohol consumption and 3) to investigate possible environments or genetic structures underlying the parallel BMI and alcohol consumption trajectories. Ultimately, our study revealed significant sex differences in the relationship between BMI change and alcohol use change. Sex-related differences were primarily observed in longitudinal weight or alcohol consumption changes, sharing common sex-specific environments with baseline. However, no differences induced by sex were found in the genetic and environmental components at baseline.

The heritability of baseline BMI (i.e., intercept) remained consistent with traditional BMI estimations (44) and was similar to those measured in the three waves 1975, 1981 and 1990 on the same cohort (h^2^ = 80% in men, h^2^ = 82% in women) (30). Estimates of the genetic and environmental components of the baseline alcohol consumption (intercept) were also consistent with the literature (41,16). Thus, these heritability estimates of BMI and alcohol consumption baseline levels were similar in men and women, and echoed those of the literature.

The phenotypic correlation between baseline alcohol consumption and change in BMI was found to be weak but significant in women. This result is consistent with large-scale studies of non-overweight women with shorter follow-ups of 7 years and 12.9 years (22-23), as these studies also showed a negative correlation between baseline alcohol consumption and weight gain in women. In our study, the non-shared environmental component of the baseline alcohol consumption and change in BMI were also significantly correlated in women, which provides additional evidence for a causal relationship, which is difficult to establish in standard epidemiological analyses.

Phenotypically, parallel changes in weight and alcohol consumption were significantly correlated in men, but not in women. This sex-specific relationship, although modestly positive in men, is consistent with what was observed in French et al. (24). The subtle positive intensity of this relationship in men was also reported in Downer et al. (21). We found a significant correlation between the non-shared environmental components of the slopes of BMI and alcohol consumption in men, demonstrating the existence of a common environmental structure influencing weight gain and change in alcohol use. This is consistent with a causal association, where increased alcohol consumption would lead to higher BMI (or vice versa). The trajectories in alcohol consumption and BMI, being consistent with the literature, reinforce both the need to appreciate the sex-induced complexity in the relationship between changes in weight and alcohol consumption, and the confidence that can be brought to the reproducibility of our twin model estimates.

A major limitation of our study lies in the statistical power in multivariate twin modeling. This lack of power mainly prevented 1) determining more precise sex differences, 2) obtaining narrower confidence intervals for heritability estimates, and 3) identifying common environments or genetics between growth factors. This last point applies particularly to men, for whom the difference in the bounds of the confidence intervals was sometimes substantial. As such, the significant phenotypic correlation between BMI and alcohol consumption intercepts in men is neither explained by a genetic correlation nor by a correlation between environmental components. The nature of this relationship thus remains unclear in our study despite some evidence of genetic structure in the literature (15). Further investigation, incorporating a larger sample size, may better capture the essence of this phenotypic relationship. However, comparable datasets are rare.

Although we found no genetic correlation between baseline BMI and alcohol consumption, a genetic correlation associating baseline BMI with a change in alcohol consumption was observed in both men and women. Such a genetic correlation seems to have not yet been observed in the literature. This genetic relationship, although weakly negative, extends insights gained in cross-sectional designs (14-15) to longitudinal settings. The nature of this genetic relationship remains undetermined, but could possibly be related to smoking, which is known to be causally associated with obesity (45-46) and alcohol consumption (14-15). Expanded research on this topic could further investigate the genetic relationship between BMI, weight change and longitudinal drinking behavior.

Our study drew strength from the 36-year follow-up time in the Finnish Twin Cohort study. Modeling the trajectory of alcohol consumption using growth factors allowed us to capture the effects associated with Finnish policies on alcohol consumption, such as those initiated in the second half of the 1980s (47), also visible in Table 1. These relatively abrupt changes in alcohol consumption provided an opportunity to detect associations between changes in alcohol consumption and weight gain at both population and individual levels. However, accordingly, linear modeling of the alcohol consumption trajectory remains questionable and simplistic in such a context. The limited number of measures of alcohol consumption to four is a constraint to quadratic modeling, although it may provide interesting results at the population level. The use of quadratic or cross-lagged (48) modeling in the study of two parallel trajectories to draw relational conclusions at the individual level may be complex in its interpretation, but remains a credible avenue for improvement.

In sum, changes in alcohol consumption and BMI were conditioned by both genetic and environmental factors, with often different intensity according to sex. In particular, the longitudinal relationship between BMI and alcohol consumption was rooted in a significant correlation between associated environmental components in men. BMI change in women was also associated with baseline alcohol consumption, as their respective non-shared environmental components were significantly correlated. A genetic correlation between baseline BMI and change in alcohol consumption was observed, however, in both men and women. Since changes in weight and alcohol consumption are behavioral traits, large disparities between cohorts can be anticipated and more studies are needed to provide an accurate and generalized relational picture. Other stratifications in addition to sex may be considered, since the correlations between indicators of obesity and alcohol consumption differ according to whether alcohol consumption is heavy or moderate (10,21). A funnel approach could, in this sense, allow a better understanding of the mechanisms associating weight change with alcohol consumption.

## Data Availability

The Finnish Twin Cohort (FTC) data is not publicly available due to the restrictions of informed consent. However, the FTC data is available through the Institute for Molecular Medicine Finland (FIMM) Data Access Committee (DAC) for authorized researchers who have IRB/ethics approval and an institutionally approved study plan. To ensure the protection of privacy and compliance with national data protection legislation, a data use/transfer agreement is needed, the content and specific clauses of which will depend on the nature of the requested data.

## Acknowledgements

The authors gratefully acknowledge Alyce Whipp of the University of Helsinki Language Service for language revision of this paper.

## Data Availability Statement

The FTC data is not publicly available due to the restrictions of informed consent. However, the FTC data is available through the Institute for Molecular Medicine Finland (FIMM) Data Access Committee (DAC) for authorized researchers who have IRB/ethics approval and an institutionally approved study plan. To ensure the protection of privacy and compliance with national data protection legislation, a data use/transfer agreement is needed, the content and specific clauses of which will depend on the nature of the requested data.

## Author contributions

The authors’ contributions to the completion of this study are divided as follows: JK and GD conceptualized the study; GD conducted the study and performed the formal statistical analyses with support from KS and JK in twin modeling; GD, KS, AL, and JK participated in the interpretation of the results and their relation to the literature; GD drafted the first version of the manuscript; All authors participated in the revision and writing of the final version of the manuscript, and approved it.

