## Supplementary Document for "Genetic and environmental factors underlying parallel changes in body mass index and alcohol consumption: a 36-year longitudinal study of adult twins"

*Drouard et al.*

March 2022

# –S1–
### Phenotypic correlations and adjustment of growth factors by covariates

### 1 Forewords

In parallel with the study of BMI and alcohol consumption trajectories for which the as sociated measures were not adjusted, a sensitivity analysis incorporating covariates was performed. The objectives were to 1) compare the effect of covariates on phenotypic correlations with those obtained in the study, 2) further refine estimates of growth factor relationships not explained by multiple covariates.

### 2 Materials & Methods

#### 2.1 Materials

A panel of covariates closely related to BMI and alcohol consumption was selected: some were time-varying such as smoking status (categories: current smoker/ former smoker/ occasional smoker/ never smoked/ uncertain) (1), age and marital status (categories: cohabiting *i.e.* married, remarried or cohabiting/ not cohabiting *i.e.* single, divorced or widowed) while education level expressed in years of study (categories: 6years or less/ from 7 to 11 years/ high school/ university/ others) (2-3) was used as a time-independent variable. The age distribution is presented in Table 1 of the paper. The education level, independent of time, is presented in Table S1. The headcounts of each class for the time-varying covariates are presented in Table S2.

**Table S1:** Description of education level in men and women. M: Men. W: Women. %M: proportion of men to total men (sample size). %W: proportion of women to total women (sample size). %T: proportion of participants (men and women) to total participants (sample size).

| Class | Men (%M) | Women (%W) | Total (%T) |
|---|---|---|---|
| 6 years or less | 449 (23.7%) | 630 (24.6%) | 1079 (24.2%) |
| from 7 to 11 years | 956 (50.4%) | 1 282 (50.0%) | 2 238 (50.2%) |
| high school | 213 (11.2%) | 454 (17.7%) | 667 (15.0%) |
| university | 174 (9.2%) | 166 (6.5%) | 340 (7.6%) |
| others | 104 (5.5%) | 33 (1.3%) | 137 (3.1%) |
| sample size | 1896 (-) | 2565 (-) | 4461 (-) |

#### 2.2 Methods

The categorical covariates with more than two categories (m) were transformed into dummy variables, i.e. $m - 1$ binary variables, to comply with the requirements of the R package used in the LGCM (*lavaan*). The time-varying covariates smoker status and marital status were used to control for measures of BMI and alcohol consumption. Education level was used to control for growth factors, *i.e.*, intercepts and slopes. This design is referred to below as the fully-adjusted design. Growth factors were provided with 95% confidence intervals.

**Table S2:** Description of time-varying covariates across the four waves of questionnaires.

| | smoking status | | marital status | |
|---|---|---|---|---|
| Questionnaire | Men | Women | Men | Women |
| 1975 | current: 725<br>occasional: 88<br>never smoker: 713<br>former: 366<br>uncertain: 4 | current: 791<br>occasional: 74<br>never smoker: 1 331<br>former: 367<br>uncertain: 2 | cohabiting: 752<br>not cohabiting: 1 144 | cohabiting: 1 151<br>not cohabiting: 1 414 |
| 1981 | current: 660<br>occasional: 84<br>never smoker: 673<br>former: 472<br>uncertain: 7 | current: 615<br>occasional: 66<br>never smoker: 1 299<br>former : 553<br>uncertain: 32 | cohabiting: 1 305<br>not cohabiting: 591 | cohabiting: 1 871<br>not cohabiting: 694 |
| 1990 | current: 581<br>occasional: 80<br>never smoker: 673<br>former: 538<br>uncertain: 24 | current: 604<br>occasional: 86<br>never smoker: 1 281<br>former: 572<br>uncertain: 22 | cohabiting: 1548<br>not cohabiting: 348 | cohabiting: 2 013<br>not cohabiting: 552 |
| 2011 | current: 340<br>occasional: 123<br>never smoker: 692<br>former: 714<br>uncertain: 27 | current: 414<br>occasional: 158<br>never smoker: 1 329<br>former: 649<br>uncertain: 15 | cohabiting: 1487<br>not cohabiting: 409 | cohabiting: 1 738<br>not cohabiting: 827 |

### 3 Results

The results obtained once fully-adjusted are presented in Table S3. The phenotypic correlations are close to those of the main study, despite one important exception: the BMI and alcohol consumption intercepts are no longer correlated in women once adjusted by covariates.

**Table S3:** Phenotypic correlations of growth factors fully-adjusted by covariates smoking status, marital status,and education level. M: Men. W: Women.

| | | Phenotypic correlations | | | |
|---|---|---|---|---|---|
| | | Intercept BMI | Slope BMI | Intercept Alc | Slope Alc |
| M | Intercept BMI | 1 | | | |
| | Slope BMI | -0.10 (-0.14, -0.05) | 1 | | |
| | Intercept Alc | 0.13 (0.08, 0.17) | -0.05 (-0.09, -0.00) | 1 | |
| | Slope Alc | -0.18 (-0.23, -0.14) | 0.17 (0.13, 0.22) | -0.52 (-0.55, -0.49) | 1 |
| W | Intercept BMI | 1 | | | |
| | Slope BMI | 0.09 (0.05, 0.12) | 1 | | |
| | Intercept Alc | -0.03 (-0.07, 0.01) | -0.05 (-0.09, -0.01) | 1 | |
| | Slope Alc | -0.09 (-0.13, -0.06) | -0.02 (-0.05, 0.01) | -0.31 (-0.34, -0.27) | 1 |

# –S2–
### Population-wide trajectories and distribution of growth values

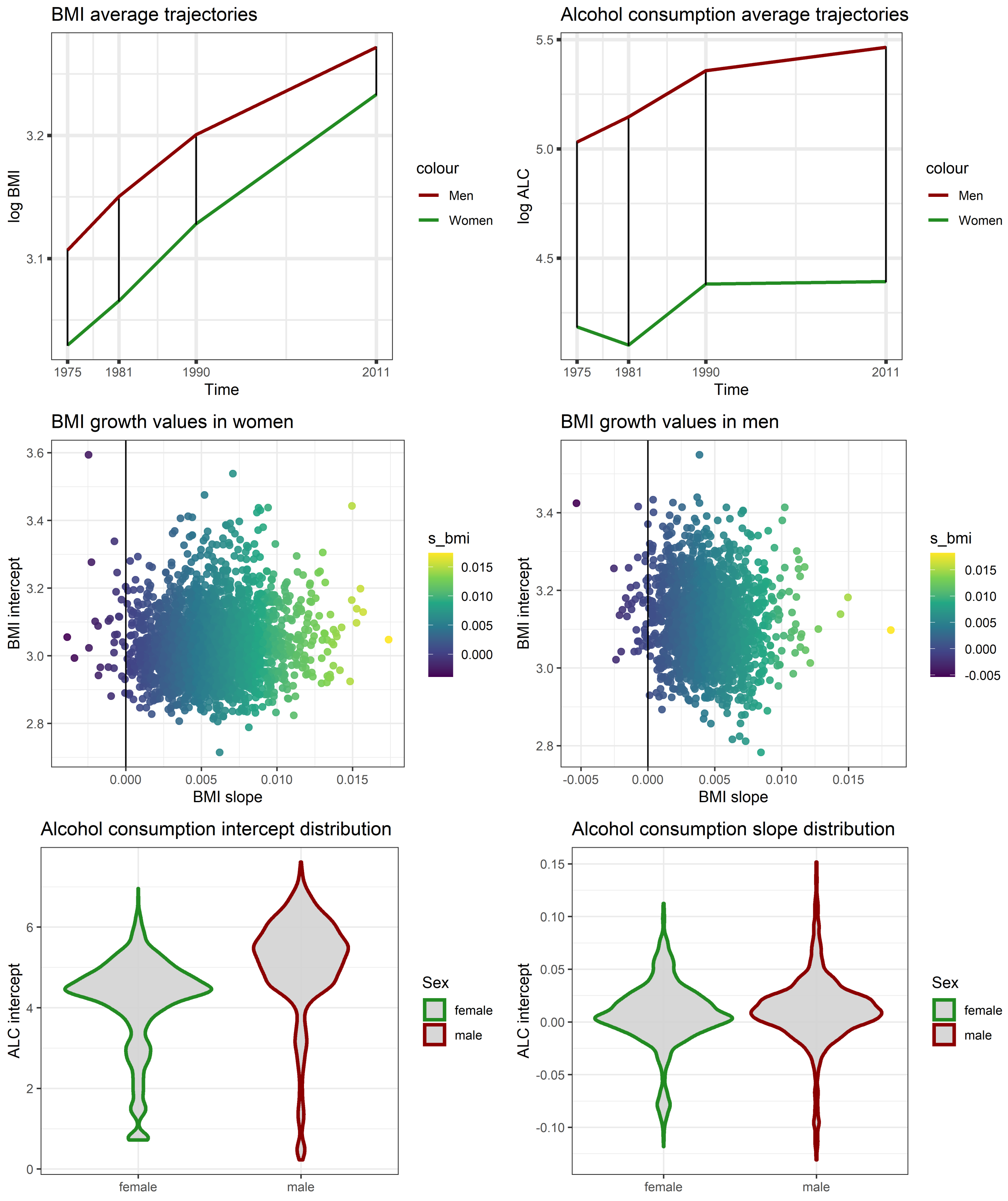


**Figure S1:** Population-wide average trajectories of log BMI and log alcohol consumption over the entire period (above), growth values of BMI growth factors (middle), and distribution of growth values for alcohol consumption (below).
